# Stability and Volatility of Human Rest-Activity Rhythms: Insights from Very Long Actograms (VLAs)

**DOI:** 10.1101/2024.01.22.24301243

**Authors:** Nandani Adhyapak, Mark A. Abboud, Pallavi S.K. Rao, Ananya Kar, Emmanuel Mignot, Gianluigi Delucca, Stephen F. Smagula, Vaishnav Krishnan

**Author notes:** Corresponding Author One Baylor Plaza St, Neurosensory BCM: MS NB302, Houston, TX 77030. Denotes Equal Contribution (Nandani Adhyapak, Mark A. Abboud).

## Abstract

**Importance:** Wrist-worn activity monitors provide biomarkers of health by non-obtrusively measuring the timing and amount of rest and physical activity (rest-activity rhythms, RARs). The morphology and robustness of RARs vary by age, gender, and sociodemographic factors, and are perturbed in various chronic illnesses. However, these are cross-sectionally derived associations from recordings lasting 4-10 days, providing little insights into how RARs vary with time.

**Objective:** To describe how RAR parameters can vary or evolve with time (∼months).

**Design, Setting and Participants:** 48 very long actograms (“VLAs”, ≥90 days in duration) were identified from subjects enrolled in the STAGES (Stanford Technology, Analytics and Genomics in Sleep) study, a prospective cross-sectional, multisite assessment of individuals > 13 years of age that required diagnostic polysomnography to address a sleep complaint. A single 3-year long VLA (author GD) is also described.

**Exposures/Intervention:** None planned.

**Main Outcomes and Measures:** For each VLA, we assessed the following parameters in 14-day windows: circadian/ultradian spectrum, pseudo-F statistic (“F”), cosinor amplitude, intradaily variability, interdaily stability, acrophase and estimates of “sleep” and non-wearing.

**Results:** Included STAGES subjects (n = 48, 30 female) had a median age of 51, BMI of 29.4kg/m^2^, Epworth Sleepiness Scale score (ESS) of 10/24 and a median recording duration of 120 days. We observed marked within-subject undulations in all six RAR parameters, with many subjects displaying ultradian rhythms of activity that waxed and waned in intensity. When appraised at the group level (nomothetic), averaged RAR parameters remained remarkably stable over a ∼4 month recording period. Cohort-level deficits in average RAR robustness associated with unemployment or high BMI (>29.4) also remained stable over time.

**Conclusions and Relevance:** Through an exemplary set of months-long wrist actigraphy recordings, this study quantitatively depicts the longitudinal stability and dynamic range of human rest-activity rhythms. We propose that continuous and long-term actigraphy may have broad potential as a holistic, transdiagnostic and ecologically valid monitoring biomarker of changes in chronobiological health. Prospective recordings from willing subjects will be necessary to precisely define contexts of use.

## Introduction

Digital health data obtained from wearable devices currently informs medical decision making in diabetes mellitus (glucose monitors), atrial fibrillation (ECG monitors) and drug-refractory epilepsy (responsive neurostimulation). The endpoints assayed by these technologies provide monitoring biomarkers^1^ to appraise treatment response and detect (and potentially predict) adverse events. When assessed remotely^2^, these measures can provide objective real time correlates of disease worsening^3,4^ and opportunities to prevent hospitalization^5^. Actigraphy, the visualization and analysis of a subject’s rest and activity patterns, offers a powerful stream of continuous data that can be collected noninvasively through limb- or trunk-worn accelerometers. Wrist actigraphy has made significant inroads in sleep medicine, offering a cost-effective and ecologically valid approach to monitor subjects with insomnia, narcolepsy, or circadian rhythm disorders^6^. Actigraphic estimates of sleep quality and quantity have been employed as endpoints in small clinical trials of pharmaceutical^7^ and behavioral interventions^8^ for sleep disorders. Wrist actigraphy has also been popularly embraced within consumer wearable devices/fitness trackers, providing personalized feedback and goals about activity levels (e.g., “step counts”) and sedentariness^9,10^.

Separate from sleep and steps, continuously worn wrist-accelerometers can also appraise rest-activity rhythms (RARs), a term preferred over *circadian* rhythms, as RARs need not display robust circadian oscillations. By applying both parametric and nonparametric techniques to multi-day long epochs, RARs can be expressed as a set of scalars that depict the rhythm’s height (amplitude), timing (acrophase), robustness (pseudo-F statistic, or “F”) and the degree of within- and across-day irregularity (interdaily stability [IS], intradaily variability [IV], respectively)^11,12^. Through large-scale efforts to measure RARs in hundreds-thousands of community-dwelling subjects (e.g., NHANES [National Health and Nutritional Examination Survey], SOL (Study of Latinos), MESA (Multi-ethnic Study of Atherosclerosis)^13-15^], we know how these parameters vary by sex, age, BMI and active employment^13,14,16-19^. Cross-sectional case-control studies have also identified RAR abnormalities in a range of chronic illnesses^20-29^. Within neuropsychiatry, depression has received the greatest emphasis, with multiple studies linking the severity of prevalent depression symptoms to reductions in robustness/routines (low F and IS, high IV), amplitude and a delayed acrophase^12,16,30-33^. This general constellation of abnormalities has been described as a “weak RAR”^19^, and is also seen in obesity^18^, frailty^34^ and Parkinson’s disease^35^.

To define these nomothetic RAR associations, virtually all studies have utilized ∼4-10 day-long recordings^13,17,19,36^, which provide little insights into whether fluctuations in symptom burden are temporally associated with fluctuations in RAR parameters. For example, despite decades of research, we have a limited understanding^37,38^ of whether the onset of depression coincides with, precedes, or succeeds a reduction in F and/or amplitude. Similarly, RAR amplitude reductions in patients on antiseizure medication (ASM) polytherapy^16^ may reflect a potentially remediable form of psychomotor retardation (“toxicity”), or a pre-existing circadian endophenotype linked to drug-refractoriness. There have been no NHANES-scale efforts to longitudinally collect actigraphy recordings in healthy or vulnerable populations, which has limited our fundamental understanding of the dynamic range and stability/volatility of RAR parameters. This knowledge is an essential requisite for the application of wrist actigraphy as a monitoring biomarker, one that may be measured repeatedly/continuously to assess disease status and/or treatment response^1^. Important examples include many serum assays, such as prostate specific antigen levels^39^ or HIV viral load^40^. In contrast, many neuropsychiatric symptoms (e.g., pain, depressed mood, sleepiness) lack precise and objective monitoring biomarkers, and have traditionally relied on psychometric instruments that are poorly suited for frequent and repeated sampling. In this report, we utilize a set of prolonged actigraphic recordings to describe how RARs vary over time. These very long actograms (“VLAs”) were discovered serendipitously amongst recordings provided by subjects enrolled in the STAGES study (Stanford Technology, Analytics and Genomics in Sleep). To further illustrate how RARs can dynamically change, we analyze a single contemporaneously annotated 3-year long VLA provided by author GD^13,41,42^.

## Methods

Actigraphy and demographic data from all STAGES subjects were downloaded with permission from the National Sleep Research Resource (NSRR^13,14^, www.sleepdata.org). STAGES is a prospective, cross-sectional, multicenter study to (i) understand the genetic architecture of sleep, and to (ii) improve the detection, treatment, and prevention of sleep disorders. To be included, subjects had to be ≥ 13 years of age and require an in-lab sleep study. Subjects were excluded if they (i) were unable to understand or read English, (ii) were pregnant, (iii) did not have a smartphone to pair with the actigraphy device, (iv) needed a sleep study for purely treatment purposes, (v) were unwilling to complete all required study assessments, or (vi) displayed an acutely unstable medical or psychiatric condition that would impact subject safety. Enrolled subjects provided responses to a comprehensive questionnaire designed to collect information about *current* medical comorbidities, prescription medications and responses to a set of validated psychometric surveys, including the ESS (Epworth Sleepiness Scale), MEQ (Morning-Eveningness Questionnaire), PHQ-9 (Patient Health Questionnaire), GAD-7 (Generalized Anxiety Disorder Questionnaire) and FSS (Fatigue Severity Scale). Subjects were instructed to wear a Huami Arc-style actigraphy device (Huami Inc)^43^ for at least two weeks and were welcome to provide more than 2 weeks of data. From a total sample of 1881 subjects, we identified 85 VLAs containing at least 90 days of actigraphy data, from which an additional 37 VLAs were excluded due to substantial rates of nonwearing (Fig. S1). Demographic features of included subjects are summarized in Fig. 3A. Examples of actigraphy recordings from individual subjects are shown in Figures 1 and 2. To preserve their anonymity, ages are provided in non-overlapping 5-year bands/ranges (e.g., 56-60, 61-65, 66-70, etc.) Actigraphy in Figure 5 (author GD) was collected through a Motionwatch 8 device (CamNtech Ltd.) in 1s epochs, which were downsampled to 60s epochs to align with STAGES analysis. The first year of author GD’s actigraphy data is publicly available at the NSRR^13,14^.

**Figure 1.**
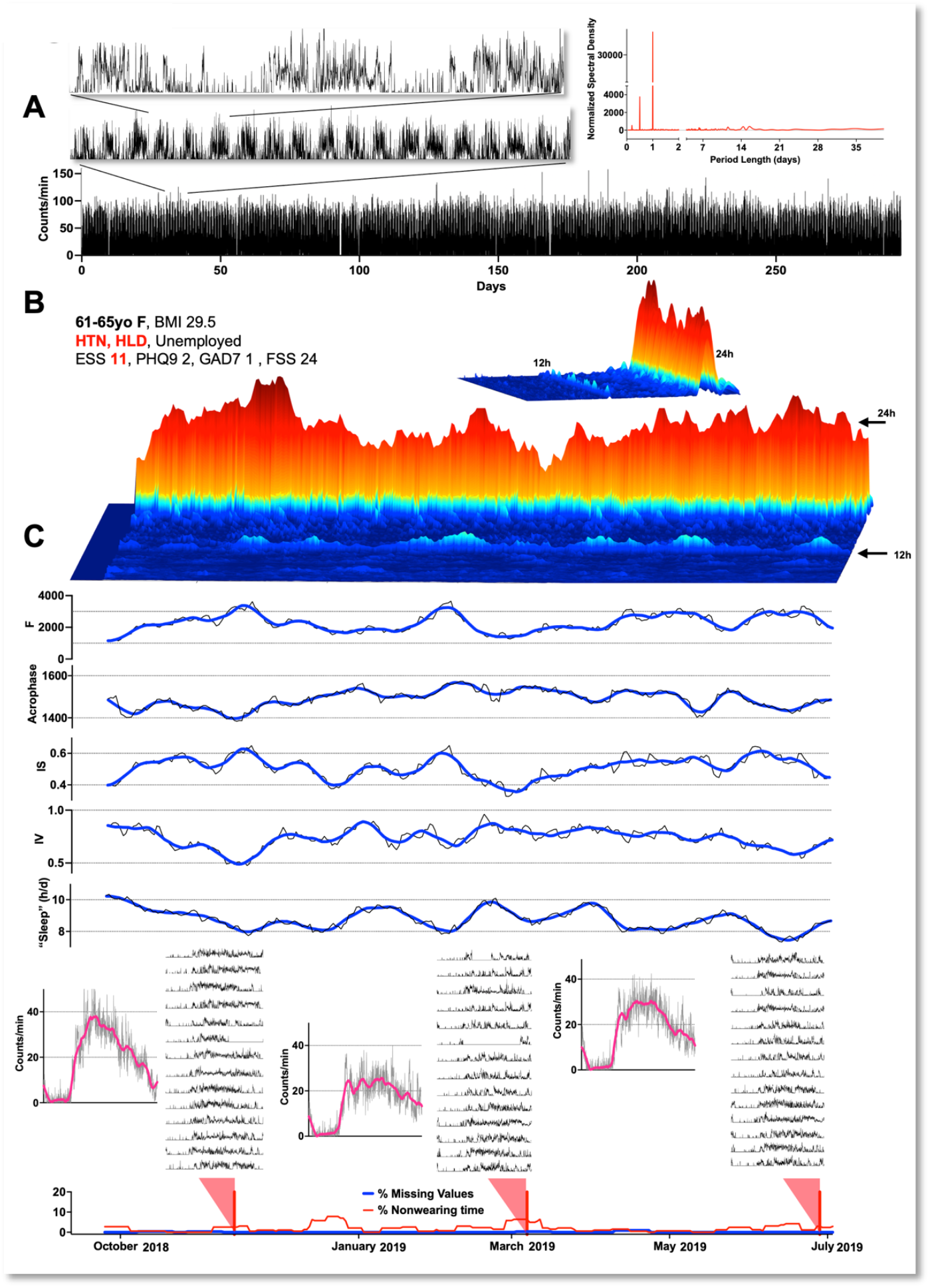
Fluctuations in RAR parameters within a 294-day long VLA (very long actogram). A: Actigraphy data can be visualized across multiple time scales, and (RIGHT) can be spectrally decomposed into constitutive frequencies. B: Three-dimensional spectrogram capturing the power of oscillations with period lengths between 2h and 28h using overlapping 14-day windows. C: Variations in RAR parameters for the same 14-day windows (grey), and smoothed (blue). BMI (body mass index), HTN (hypertension), HLD (hyperlipidemia), ESS (Epworth Sleepiness Scale), PHQ9 (9 item Patient Health Questionnaire), GAD7 (7 item Generalized Anxiety Disorder Scale), FSS (Fatigue Severity Scale).

**Figure 2.**
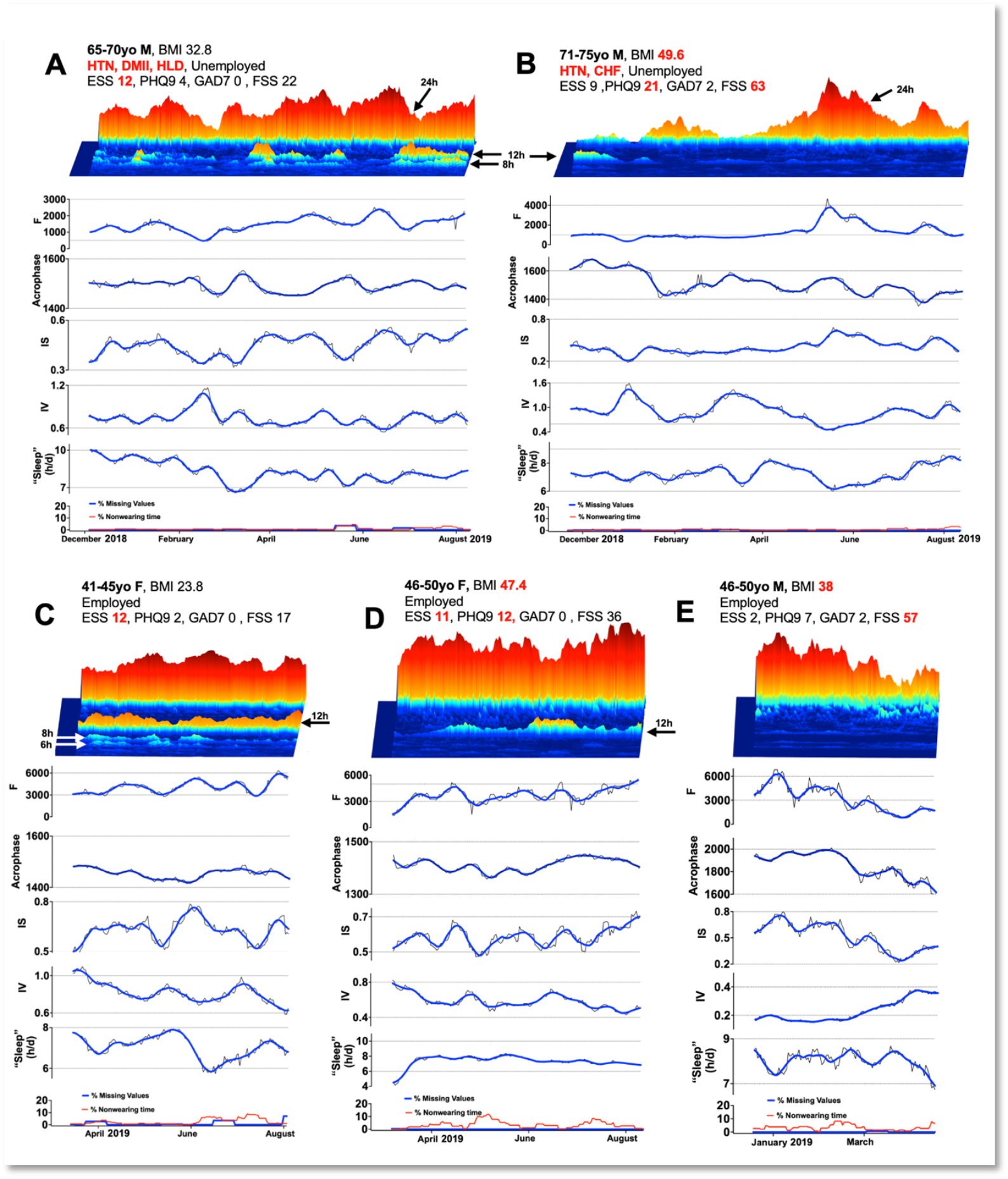
Other representative VLAs. A: This VLA captures well-formed ultradian rhythms of activity with period lengths of 12 and 8h. Estimates of total sleep time varied between 7-10h/d. B: In this subject, poor circadian rhythmicity at the start of the recording associated with a relatively delayed acrophase and low F, all of which appeared to resolve over the course of 5-6 months. C: 6, 8 and 12h ultradian rhythms were observed in this VLA. D: Another exemplary VLA demonstrating a transient boost in semicircadian power occurring in concert with a transient reduction in circadian power. E: VLA demonstrating a gradual reduction in circadian power over time, associated with an acrophase advance. Each graph depicts both raw RAR parameters in 14-day windows (grey) and smoothed (blue). HTN (hypertension), HLD (hyperlipidemia) DMII (Type 2 diabetes mellitus), CHF (congestive heart failure).

**Figure 3.**
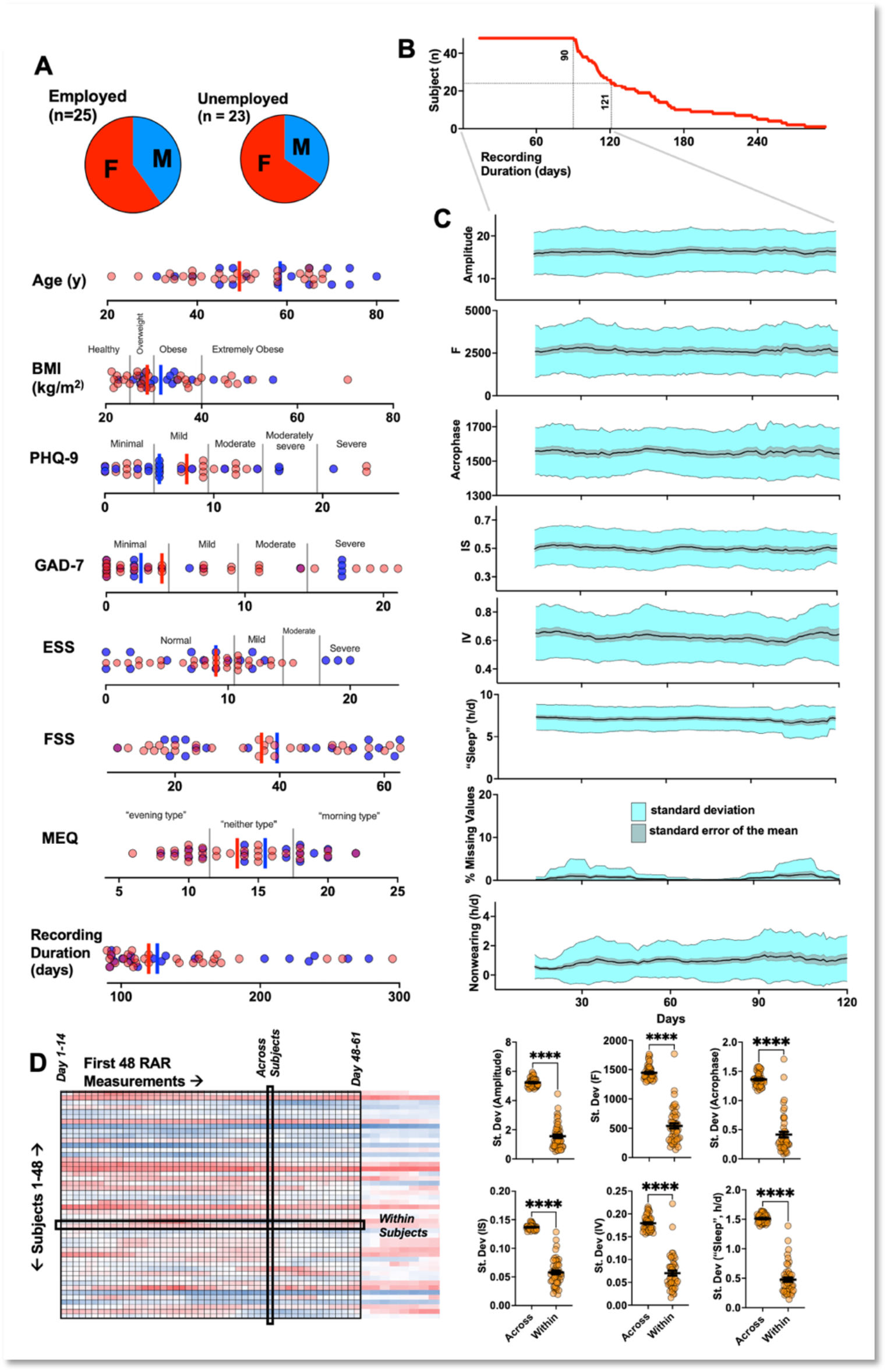
RAR parameters averaged across the cohort. A: Demographic and psychometric data from selected STAGES subjects. Vertical bars depict median (blue – male, red-female). B. 48 subjects provided at least 90 days of recording, and 24 subjects provided at least 121 days of recording. C: RAR parameters averaged over time, by aligning start dates of recordings, depicting standard deviation and standard errors of the mean for all parameters. D: Over the first 48 daily actigraphic parameters (summarizing 61 days [48+13]) across all 48 subjects, within-subject standard deviations were approximately a third of across-subject standard deviations. ^****^ denotes p <0.0001.

From all raw actograms, partial-day *tail* recordings were discarded, resulting in an integer number of full “days” (defined as midnight to midnight). Actograms lasting *n* days were transformed into a matrix containing 20160 rows (14 days *x* 1440 rows/days) and *n-13* columns, summarizing days 1-14, days 2-15, and so on. Each column of data was spectrally decomposed through the Lomb Scargle periodogram^16,44^ over a discrete set of period lengths (MATLAB). RAR parameters for each 14-day window were individually quantified as described previously (github.com/JessLGraves/RAR)^16,33,45^, regressing log-transformed actograms to a sigmoidally transformed cosine curve, generating measures of RAR robustness (“pseudo”-F statistic, or simply “F”) and acrophase. Amplitude measurements were derived from a standard cosinor regression^46^. Nonparametric measures (intradaily variability [IV] and interdaily stability [IS])^11^ were calculated using github.com/wadpac/GGIR. Epochs of nonwearing were defined as bouts of contiguous zero activity lasting ≥ 100 minutes. In the absence of detailed sleep diaries over these extended recordings^47^, we estimated the occurrence of “sleep” bouts as epochs containing between 4-99 minutes of contiguous zero activity^16^, providing a measure of mean daily total sleep times over 14d epochs.

Spectrograms (shown as surface plots) were plotted using the Matlab *surf* function. All other graphs and statistical analyses were prepared using Prism GraphPad 10. A student’s T test was employed to conduct pairwise comparisons in Fig. 3D. A mixed effects model was implemented to examine group-wise variations of RAR parameters over time, using restricted maximum likelihood and the Geisser-Greenhouse correction (Fig. 4). Changes in subject-specific RAR parameters over time (Fig. 1 and 2) were smoothed with a 2^nd^ order polynomial averaging 10 nearest neighbors. Multiple linear regression was employed to determine the odds ratios with which age, female sex, BMI, employment status and psychometric scores at consent (ESS, PHQ-9, GAD7, MEQ and FSS) impacted within subject standard deviations in RAR parameters calculated over the first 90 days of recording (Fig. S2B). For all models, variance inflation factors for all parameters were < 4.0.

**Figure 4.**
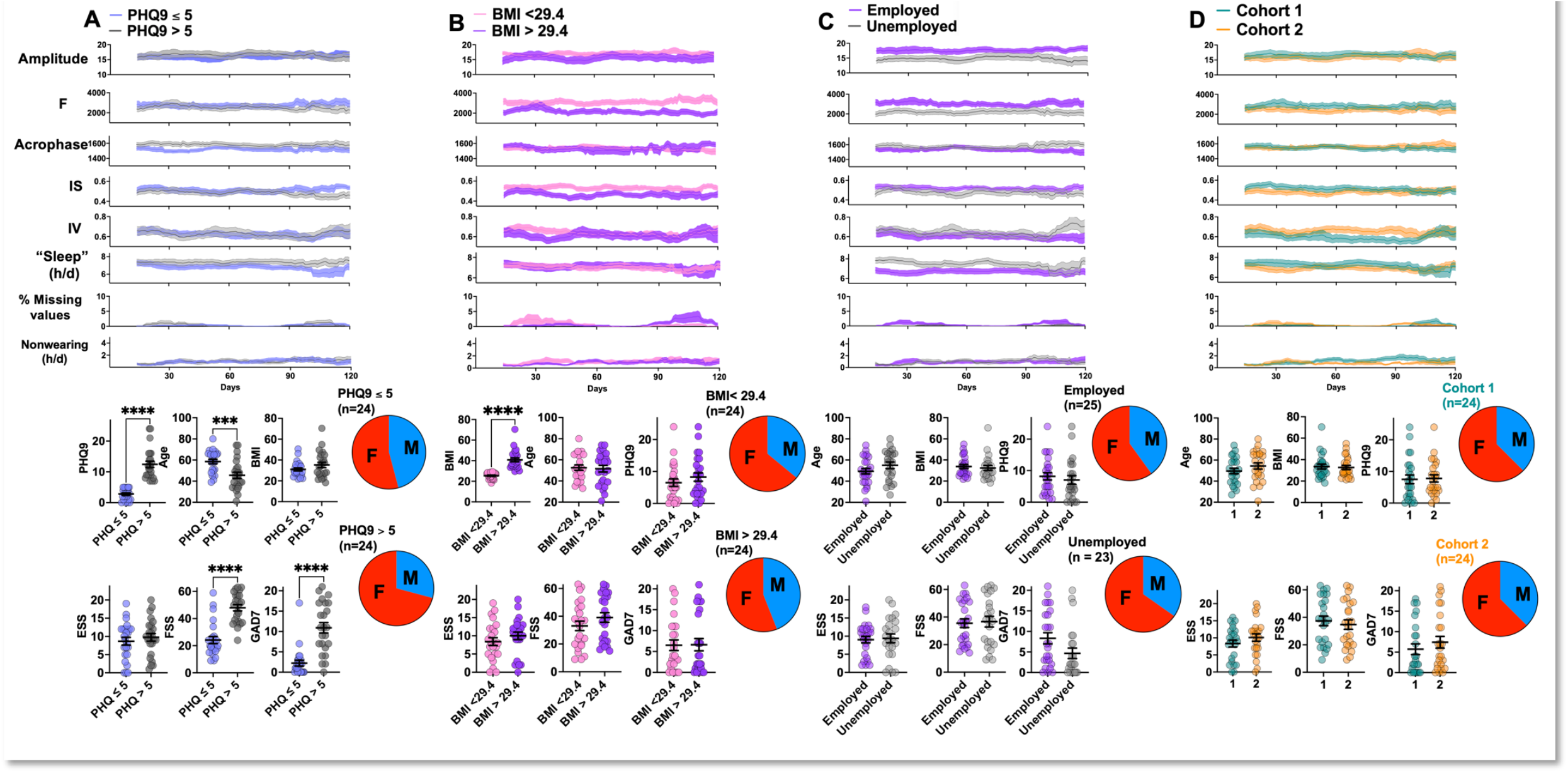
RAR stability in demographically or psychometrically defined cohorts of subjects. A. Subjects with higher (than median) PHQ-9 scores were also younger, more likely to be of female sex and displayed significantly higher GAD7 and FSS scores. Subjects with BMI > 29.4 (B) and those that were unemployed (C) displayed reductions in F that were stable across the recording duration. D: 48 subjects were manually divided into matched groups, simulating a protracted “baseline” observations period prior to treatment or placebo exposure. ^***, ****^ denote p<0.001, 0.0001 respectively.

## Results

From a total of 85 VLAs that contained at least 90 days of actigraphy data, we excluded recordings from 37 subjects with substantial nonwearing, particularly within the first 30 days (Fig. S1). Compared against the remainder of all STAGES enrollees, our 48 included subjects were older (odds ratio 1.03 [1.01 to 1.06]) and displayed lower MEQ scores (odds ratio 0.91 [0.84 to 0.96]), but were not significantly different in sex, BMI or ESS, FSS, PHQ-9 and GAD-7 scores. Fig.1 illustrates an exemplary VLA from a 61-65yo female subject that provided a 294-day long recording. At consent, the subject was unemployed, reported comorbidities of hypertension and hyperlipidemia, and was taking gabapentin, atorvastatin, losartan and melatonin. She displayed psychometric evidence of excessive subjective sleepiness (ESS ≥ 9) but low/normal levels of prevalent depression (PHQ9), anxiety (GAD7) and fatigue symptoms (FSS). As shown in Fig. 1A, circadian rhythmicity is difficult to discern from a compressed raw actogram of this length. When spectrally decomposed, prominent peaks of rhythmicity were observed at period lengths corresponding to the circadian oscillator (1 day),12 hours and 4.8 hours. Well-defined infradian peaks (e.g., 7 days or 28 days) were not observed in this subject.

To observe how specific RAR parameters varied as a function of time, we divided the recording into overlapping 14-day epochs (e.g., days 1-14, days 2-15, etc.) Fig. 1B depicts the spectral features of these 14-day epochs as a surface plot, showing marked fluctuations in the power/amplitude of the circadian oscillator over time. Variations in the power of 12h-long rhythms (*circasemidien* or *semicircadian*^48,49^) were also observed, occurring largely independent of the circadian power. Using the same 14-day windows, we also examined how a set of RAR parameters evolved over time (Fig. 1C). “F” scores (or the “pseudo-F statistic”) displayed up to three-fold variation (ranging from 1000-3000), mirroring changes in circadian amplitude. Acrophase also vacillated between 1400-1600. Among nonparametric RAR measures^11^, we observed considerable fluctuations in both IS (stability across days) and IV (measuring rhythm fragmentation). IS strongly correlated with F and was correlated inversely with IV. Averaged total daily “sleep” times (see Methods) ranged between ∼8-10h/d. While the burden of missing values was low, estimates of average watch nonwearing measured as high as 10% (∼2.4h/d).

We conducted a similar analysis for the remaining 47 VLAs and highlight some representative trends/motifs in Fig. 2. Approximately 8 months of data was provided by a 66-70yo male subject with excessive sleepiness (Fig. 2A), who displayed prominent 12- and 8h-long ultradian rhythms of activity. Estimates of total “sleep” time varied between 7-10h/d and could not be explained by nonwearing. Fig. 2B summarizes another ∼8-month long recording from a 71-75yo morbidly obese unemployed man with hypertension and congestive heart failure, who reported high rates of fatigue and PHQ-9 scores compatible with severe depression. At the start of the recording, this subject’s circadian and semicircadian peaks of spectral power were remarkably similar in amplitude. Over time (or secondary to unknown treatment), the subject demonstrated a gradual improvement in circadian power, an acrophase advance and an improvement in total “sleep” times. The subject in Fig. 2C provided a ∼5-month long VLA with a rich constellation of 12-, 8- and 6h long ultradian rhythms of activity. Fig. 2D depicts a ∼5-month long VLA from a morbidly obese 46-50yo employed woman with PHQ-9 scores compatible with moderate depression. This recording captured a transient boost in semicircadian power associated with a corresponding transient reduction in circadian power. And finally, Fig. 2E shows a ∼4 month-long VLA from a 46-50yo employed obese woman with particularly high ratings of fatigue. In this example, we observed a gradual reduction in F over time, associated with an expected increased IV and decreased IS. This record did not feature any well-defined ultradian peaks. Together, these examples illustrate how measures of RAR morphology, robustness/stability and “sleep” calculated during the first 14-day epoch varied substantially over subsequent weeks/months.

Next, we shifted our focus to examine how RAR parameters behaved when averaged across the cohort. 30/48 subjects were female and 25/48 were employed (Fig. 3A). Overall, subjects displayed a median age of 51, a median BMI of 29.4kg/m^2^ and high median ESS scores (10, Fig. 3A). All subjects provided at least 90 days of recording (defining their inclusion), and 24 subjects provided greater than 121 days of recording (Fig. 3B). In Fig. 3C, we tallied the mean and variance of observed RAR parameters over time. In contrast to the dynamic changes observed within individual subjects, group-level RAR parameters remained relatively stable over the first 120 days of recording, including days 90-120, where the overall standard deviation (SD) remained stable despite the sample size dwindling by ∼50%. When we aligned subject data by calendar date, we did not detect any obvious seasonal fluctuations in RAR parameters (Fig. S2A), although this analysis is limited by sub-annual recording durations and substantial variations in start dates^50^. To qualitatively assess the extent of within-subject RAR fluctuations, we compared the daily standard deviations of each RAR parameter across all 48 subjects to standard deviations within individual subjects across the first 48 recording epochs. On average, within-subject standard deviations were approximately a third of those measured across subjects over a single epoch (Fig. 3D). To understand whether certain demographic and psychometric characteristics impacted within-subject variability, we developed independent multiple linear regression models for each RAR (Fig. S2B). RAR amplitudes and daily “sleep” times were more invariant in younger subjects. High ESS and PHQ9 scores (at consent) predicted greater variations in IV, while low MEQ scores (“evening” chronotypes) predicted greater stability in acrophase (Fig. S2B).

Since the PHQ-9 is a widely utilized survey of prevalent depression symptoms, we explored whether depression-related RAR changes were at all present in our cohort, and whether they remained stable over time (Fig. 4A). Using a median-split of PHQ-9 responses, we segregated a low (≤5) and high (>5) depression subgroups. Subjects in the high PHQ9 cohort were significantly younger, more likely to be female, and scored significantly higher on GAD-7 and FSS surveys. Scores of F and amplitude were similar initially and across time (potentially due to age and sex differences^19^). High PHQ9 subjects also accumulated greater total sleep time (*group* x *time*, F_106, 4490_ = 2.32, p<0.0001). Community-derived associations between obesity and RAR changes have revealed similar results, whereby high BMI scores are associated with lower amplitudes^18^ and F scores^16,18^. Using a similar median split, we found that F scores in high BMI subjects were consistently and significantly elevated throughout the recording period (*group* x *time*, F_106, 4390_ = 1.95, p<0.0001, Fig. 4B). We also examined associations with employment status: unemployed individuals tend to display lower amplitudes, acrophase delay, lower F and higher total “sleep” times^16^. All four of these relationships were observed in unemployed subjects (Fig. 4C), including amplitude (*group* main effect F_1,46_ = 4.4, p<0.05), acrophase (*group* x *time* F_106,4432_ = 4.4, p<0.05), F (*group* main effect F_1,46_ = 6.33, p<0.05), total daily “sleep” (*group* x *time* F_106,4490_ = 1.4, p<0.01).

Finally, to simulate the utilization of RAR parameters as a clinical trial biomarker, we manually divided our 48 subjects into two equally sized cohorts matched in demographic and survey responses (Fig. 4D). Cohorts “1” and “2” defined in this fashion displayed similar RARs at onset, and F, amplitude and “sleep” measures remained similar over the recording duration.

In Figure 5, we illustrate a 1094-day long VLA provided by author GD^41,42^. At the start of the recording (June 2016), the subject was a 61-65yo employed man (BMI 26.3kg/m^2^) with a history of uncomplicated cholelithiasis and nephrolithiasis, taking only acetaminophen for mild arthritis-related pain as needed. Two days after a 2016 Christmas day lunch with his family, he (and several members of his family) fell sick with a particularly severe case of the flu, requiring several days of bedrest. This incident was associated with a relatively steep but transient drop in circadian power and F. A second health issue occurred in 2018, when the subject experienced retinal detachment with hemorrhage on March 26 and March 28. Given the severity of hemorrhage, the subject was instructed to maintain total bed rest until surgery could be performed (April 24, 2018). More limited bedrest was required for the next several months, as the subject underwent repeated rounds of laser therapy. At around the onset of his symptoms in March, we observed an expected reduction in circadian power together with a notable patch of ultradian silence. In July of that year, a more protracted reduction in circadian power was accompanied by a boost in semicircadian power, with stable F scores. Clinically, this period was associated with a reinstitution of his daily walks.

**Figure 5.**
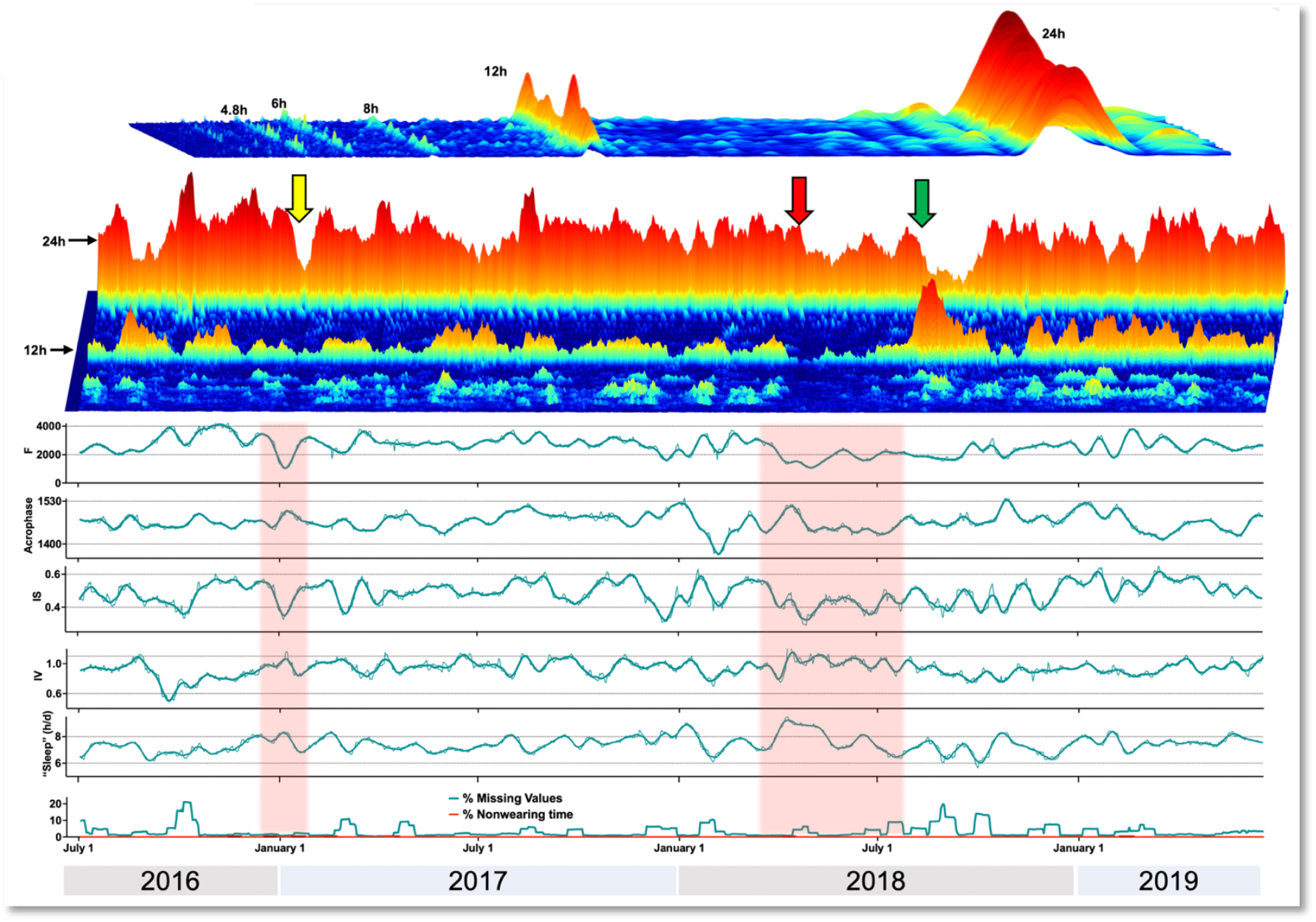
Fluctuations in RAR parameters within a 1094-day long VLA (very long actogram). Yellow arrow: depicting a steep but transient drop in F, amplitude, IS associated with a case of the flu. Red arrow: Patient experiences two retinal hemorrhages in two days, followed by strict bed rest for several weeks. This is associated with a step wise decline in amplitude, F and “sleep”. Green arrow: A boost in semicircadian power is observed as the patient resumes his normal activity levels, associated with a gradual restoration of circadian power by October.

## Discussion

As quantitative biomarkers of chronobiological health, RARs reflect the functional output of brain circuits subserving circadian rhythmicity, which in turn modulate (and are modulated by) closely interconnected brain networks that may be dysfunctional (e.g., stroke, epilepsy), exogenous environmental (e.g., sunlight exposure, exercise) or iatrogenic influences (e.g., sleep aids), and/or socioeconomic differences (e.g., employment, race/ethnicity). RAR differences by health-related variables may inform public health interventions designed to mitigate demographic disparities in healthy aging, morbidity and mortality^12,17-19,33,51^. By comparing RAR features between cohorts (e.g., controls vs affected) using data from days-long recordings, we have implicitly assumed that these differences remain stable over time. Our study visualizes this cohort-level stability through real-world recordings of subjects that shared a requirement for in-lab polysomnography, *and* a willingness to provide months of actigraphy data. Beyond this selection bias, our sample was quite heterogeneous in age, BMI and extent of neuropsychiatric comorbidity. We found that even in relatively small cohorts (24-48 subjects) over 90-120 day recording durations (aligning with length of many clinical trials^52-54^), averaged RAR parameters remained stable over time, as did demographically mediated differences in RAR (e.g., low F in high BMI or unemployed subjects). This stability occurred in the absence of any planned or coordinated intervention, approximating a placebo or “standard of care” cohort. We hypothesize that continuous actigraphic surveillance, applied as an adjunct clinical trial biomarker, may connect clinically meaningful improvements in wellbeing/quality of life with objective stereotyped RAR changes. In neurological conditions that feature motor decline as a core symptom, parallel efforts that longitudinally survey daytime activity *counts*^3^ and total *steps*^4^ are already underway. The simultaneous incorporation of RAR parameters may signal biomarkers of improvements in non-motor symptoms, including sleep and mood. This approach would also transparently depict the neurobehavioral “costs” of an intervention, for example, by quantifying the increased somnolence that occurs with anticonvulsant treatment, or the reductions in robustness that may associate with the depressive side effects of interferon treatment.

Simultaneously, we observed relative *volatility* in RAR parameters when appraised within individual subjects, with variations that paralleled RAR differences in well-defined demographic or health-related changes. For example, from over 12,000 7-day long recordings of subjects between 3-80 years of age (NHANES), average IS scores decreased with age (from ∼1 to ∼0.8), while IV increased with age (from ∼0.6 to ∼0.9)^19^. We observed fluctuations of a similar magnitude within individual subjects over a timescale of a few weeks (Figs. 1,2). Similarly, in a study of community dwelling women aged ≥ 65 (Study of Osteoporotic Fractures, SOF), subjects with high prevalent depression symptoms displayed a significant ∼20% reduction in F (∼740 vs ∼918 in controls)^30^. F scores in our 48 subjects displayed an average coefficient of variation of ∼27% (i.e., with a standard deviation that was 27% of the mean). As patient-driven interest in wearable technologies continues to grow, our findings provide a starting point to integrate actigraphy-derived RAR assessments routinely into the individualized management of chronic illness, beyond those that primarily impact motor function. At the very least, such RAR “dashboards” would provide objective estimates of sleep and activity levels, endpoints that are notoriously prone to response distortion. Our study illustrates one such dashboard prototype that concisely provides a top-down view of subject-specific trends in RARs that may be captured in between routine clinic follow up visits, and which may be annotated prospectively by both physicians and patients. Just as achieving specific “step count” targets may improve cardiovascular fitness^55^, we hypothesize that setting personal goals for RAR robustness and timing may positively impact mental wellbeing.

We outline several key limitations. First, compared with community-derived actigraphy efforts (e.g., NHANES, SOL, MESA, SOF), our sample is relatively small. Without event or symptom diaries, we cannot make any inferences about the whether our observed RAR fluctuations were at all linked to improvements or deteriorations of health. By virtue of our selection criteria (≥ 90-day recordings), we cannot rule out the possibility that estimates of RAR volatility within these subjects was linked to their greater motivation to engage with a research actigraphy recording. Second, like most wearable-derived data streams, our time series contained missing values and epochs of nonwearing. We elected to transparently juxtapose these epochs with RAR parameters, and rather than interpolating missing segments, we chose to minimize their acute influences on RAR parameters by adopting a wide 14-day window of analysis. This approach intentionally filters out higher frequency oscillations and focuses on underlying smooth trends in circadian behavior. We recognize that certain clinical scenarios may require wider or narrower windows of observation. Third, as with any actigraphy-derived estimation of sleep occurrence or timing, our algorithm may mis-classify epochs of quiet wakefulness. Absent sleep diaries (which are incomplete even in shorter actigraphic recordings^47^), our estimates of total “sleep” time may include naps that occur outside the main sleep period.

## Conclusions

In an exploratory analysis of months-long wrist actigraphy, we find that the morphology and robustness of rest-activity rhythms vary markedly with time when assessed within individual subjects. In contrast, when such longitudinally assessed RAR parameters are averaged across a demographically defined cohort of subjects, these biomarkers remain stable. To further define the monitoring biomarker potential of wrist actigraphy-derived RARs, we require long-term prospectively annotated recordings from willing subjects.

## Data Availability

Source data for this study are available for download with permission from the National Sleep Research Resource (www.sleepdata.org).

## Acknowledgments/Funding/Support

The STAGES study was funded by the Klarman Family Foundation. VK received support from the NIH (R01NS131399, K08NS110924) and seed funding made possible by the Baylor College of Medicine’s Office of Research. SFS receives support from the NIH (R01MH125846).

## Supplemental Data

**Supplementary Figure 1.**
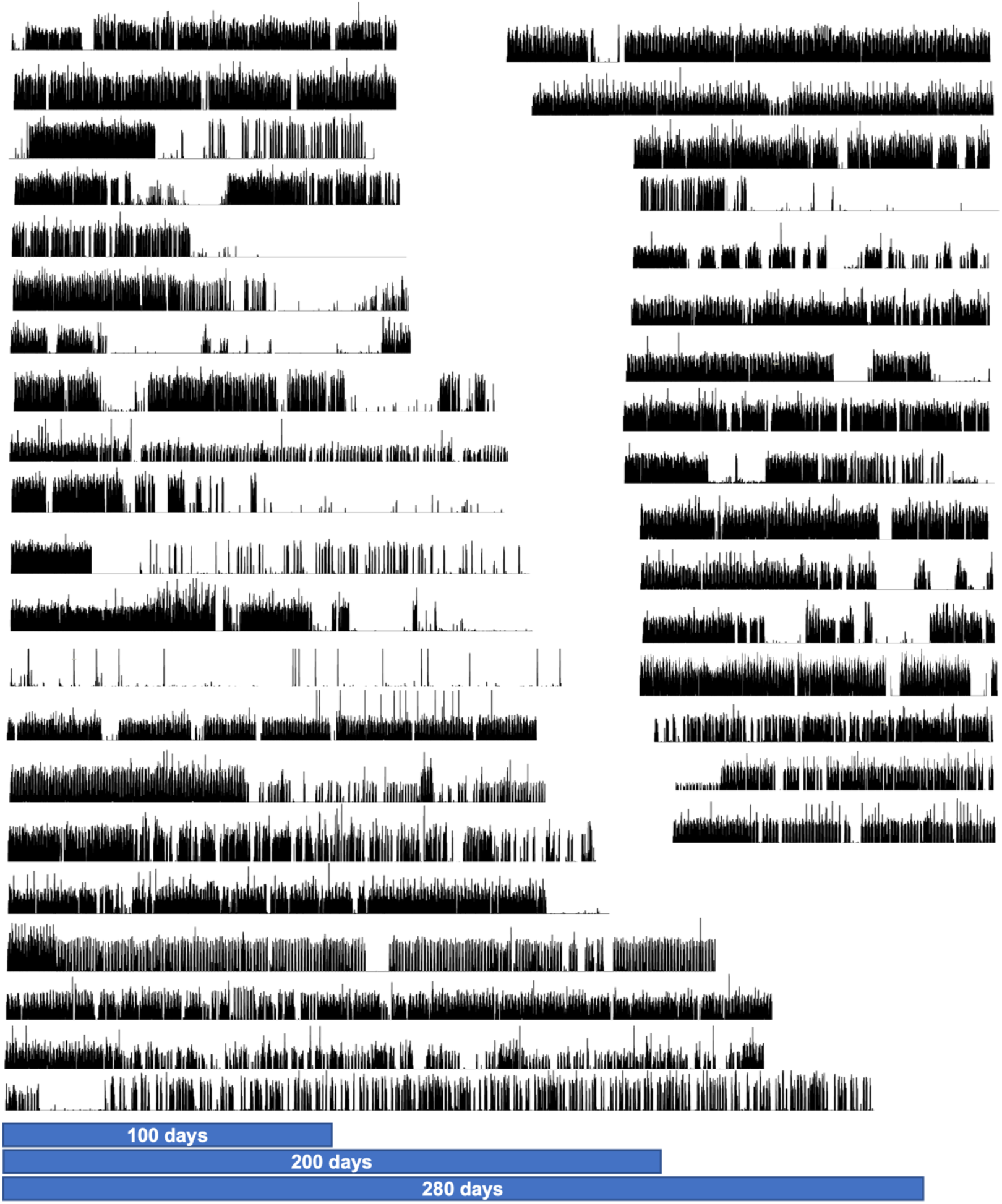
85 STAGES subjects provided actigraphic recordings ≥ 90 days, from which the above 37 were excluded due to significant nonwearing, especially when it occurred close to recording onset (when health-related data, prescribed medications and psychometric surveys were collected.

**Supplementary Figure 2.**
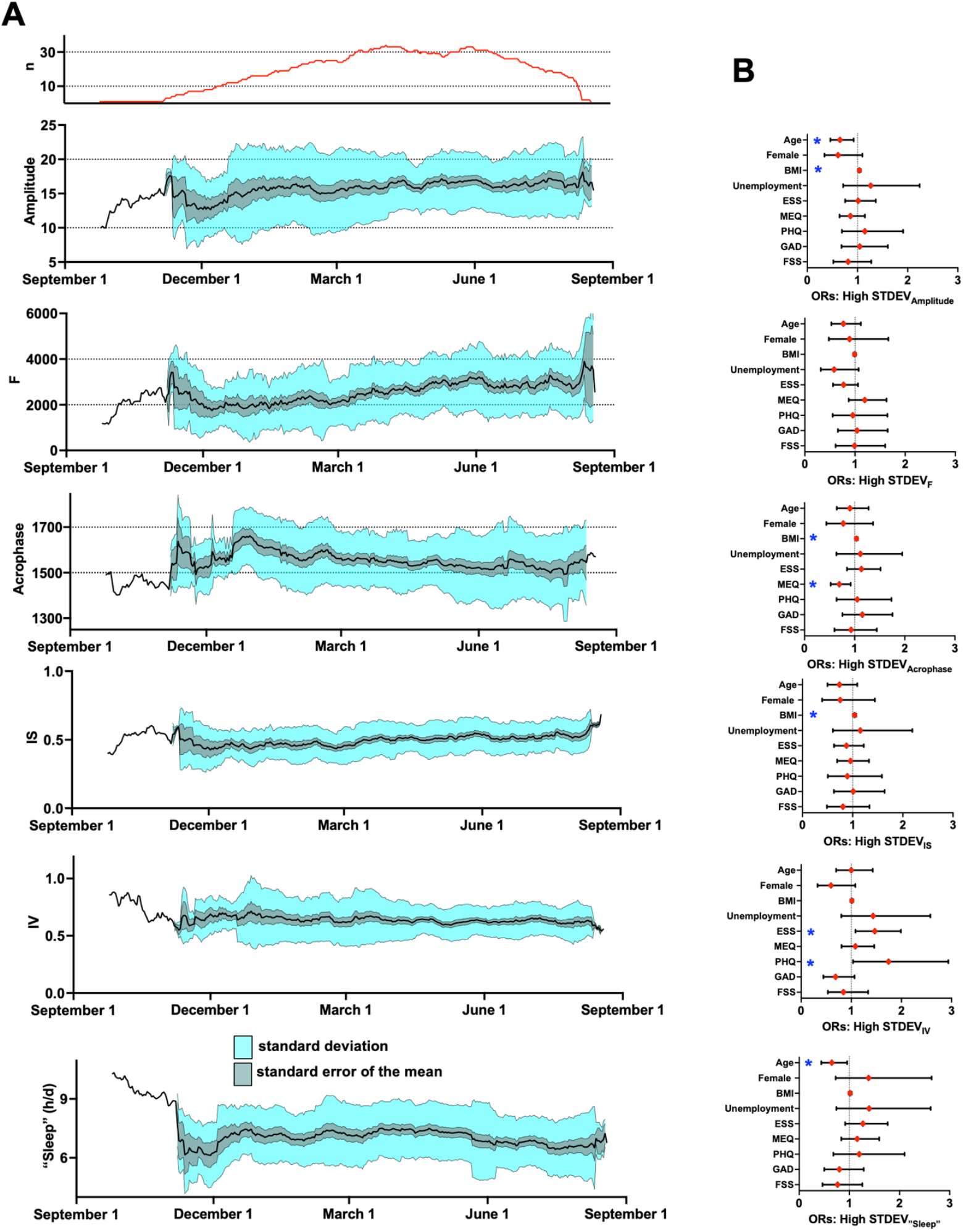
A: Mean, standard deviation and standard errors of the mean for RAR parameters aligned by calendar date. Top curve depicts real-time cohort sample size. B: Odds ratios (ORs) and 95% confidence intervals from multiple linear regression models examining the influence of age, sex, BMI, employment status and psychometric variables (ESS, PHQ9, GAD7, MEQ and FSS) on the standard deviation of RAR parameters over the first 90 days of recording. ^*^ denotes p <0.05.

